# Dysregulated B Cell Education Drives Autoimmunity and Immunodeficiency in Down Syndrome

**DOI:** 10.64898/2026.11.11.26367301

**Authors:** Meredith Ramba, Sofija Buta Panov, Haley E Randolph, O’Jay Stewart, Anass Abbad, Amy Ku, Rachel Brody, Gyu Ho Lee, Florian Krammer, Dusan Bogunovic

## Abstract

Individuals with Down syndrome (DS) present with both increased rates of autoimmunity and immunodeficiency. To understand this paradox, we first examined lymph node (LN) architecture and immune cell composition in infants with and without DS, as well as peripheral blood B cells from unrelated adults with and without DS. Most strikingly, LNs from individuals with DS exhibited a marked paucity of germinal centers compared with typical lymph nodes. Stromal cells within DS LNs expressed significantly lower levels of the adhesion molecules *ICAM1* and *VCAM1*, accompanied by significantly reduced *CXCR5* expression within B cell follicles, suggesting impaired B cell migration and follicular organization. Although fibroblastic reticular cells (FRCs) were present at comparable frequencies in DS and control LNs, MARCO lymphatic endothelial cells (MARCO-LECs), another critical stromal cell population supporting LN architecture, were significantly reduced. In addition, follicular dendritic cells (FDCs), which are essential for antigen retention and B cell selection within germinal centers, were also substantially reduced. To determine the consequences of these abnormalities on humoral immunity, we performed paired single-cell RNA sequencing and V(D)J sequencing of peripheral blood B cells from adults with and without DS. DS memory and CD86^+^ switched memory B cell populations exhibited significantly elevated reactive oxygen species and PI3K–AKT–mTOR signaling gene signatures, coupled with reduced levels of somatic hypermutation in the memory population and increase usage of the autoreactivity-associated *IGHV4-34* heavy-chain gene. Collectively, these findings suggest that early-life disruption of LN architecture, characterized by reduced expression of adhesion molecules, selective loss of MARCO-LECs, and depletion of FDCs, impairs germinal center function and B cell education. This defective microenvironment likely contributes to inadequate affinity maturation, as reflected by diminished somatic hypermutation and a more autoreactive B cell repertoire in adults with Down syndrome.

## Introduction

Down syndrome, most often genetically presenting as trisomy 21 (T21), occurs in around 1 in 700 live births.^1^ Clinically, T21 presents as a constellation of clinical features with considerable phenotypic variability inclusive of musculoskeletal, neurodevelopmental, cardiac, and respiratory disorders, as well as immune dysregulation.^2,3^ Immune dysregulation presents in both directions: 1) increased autoimmune and autoinflammatory diseases, such as thyroiditis, eczema, alopecia areata and type I diabetes, and 2) increased immune deficiency as defined by risk of severe infectious diseases.^2–5^ This defect can be driven by innate immune responses^6^ but also by inadequate adaptive immune responses, as individuals with T21 are documented to have considerable variability in response to vaccination. While not fully understood, in some cases response to immunization is decreased compared to disomic individuals (D21), while in others, antibody titers wane more quickly in T21 individuals than in their D21 counterparts.^7–10^

The observed increase in severe infectious diseases could be due to the well-described contracted B cell compartment in both children^11^ and adults with T21.^12–15^ However, this decrease in B cells is likely only a minor influence. The theoretical B cell receptor (BCR) diversity is conservatively thought to result in 10¹¹–10¹³ distinct clonotypes, while actual diversity due to typical B cell numbers in a disomic individual is around 10⁷–10⁸ distinct clonotypes.^16^ Thus, even a contracted B cell compartment would still result in 3×10^6^ to 3×10^7^ distinct naïve BCR clonotypes, likely providing sufficient coverage for infectious diseases. Adding to this notion, the documented increase in autoimmune diseases due to autoantibody production^12^ suggests that the driver of dysregulated adaptive B cell immunity likely lies elsewhere, perhaps in B cell education. Some studies have reported significantly fewer class-switched memory B cells in individuals with T21 compared to D21,^9,15,17^ with our group and others reporting significantly elevated plasmablasts in T21.^12–14^ One study documented that tonsils from children with T21 showed significantly smaller germinal centers, with significantly fewer BCL-6 positive cells, a canonical marker of germinal center B cells.^18,19^ However little is known about how germinal centers are formed, maintained, and function in T21. Perhaps even fewer details exist on the outcomes of B cell maturation, such as levels of somatic hypermutation.

In this study, we explore B cell education in individuals with T21 across age. Specifically, we characterize germinal center formation in lymph nodes of infants with T21, compared to D21 infants, and document in detail the immune and stromal cell populations of these lymph nodes using spatial transcriptomics. Furthermore, we investigate B cell maturation via somatic hypermutation and class switching in CD19+ sorted B cells from peripheral blood mononuclear cells (PBMCs) of D21 and T21 adults, using paired single-cell RNA sequencing and V(D)J sequencing, providing an in-depth investigation of the B cell compartment in T21.

## Results

### T21 lymph nodes have a paucity of germinal centers

We began by investigating lymph nodes (LNs) from infants with T21, as they are a site of B cell education and germinal center formation. To assess lymph node structure, we stained four lymph nodes from typical disomic infants (D21) and three lymph nodes from infants with T21, all within the age range of 0-4 years old, with hematoxylin and eosin (H&E) (Figure 1A - C). Clinical information from autopsies and pathology reports of these patients did not suggest lymph node or immune pathology in any of the individuals (Figure S1A). As expected, H&E staining of all D21 lymph nodes revealed a typical lymph node structure, with apparent primary B cell follicles, germinal centers (secondary follicles), and T cell zones (paracortex) in each lymph node (Figure 1B, S1B). In contrast, T21 lymph nodes appeared to lack clear germinal center formation. T21 LN follicles appeared not only scarcer, but also structurally less well-defined and more hypocellular compared to D21 lymph nodes (Figure 1C, S1C).

**Figure 1.**
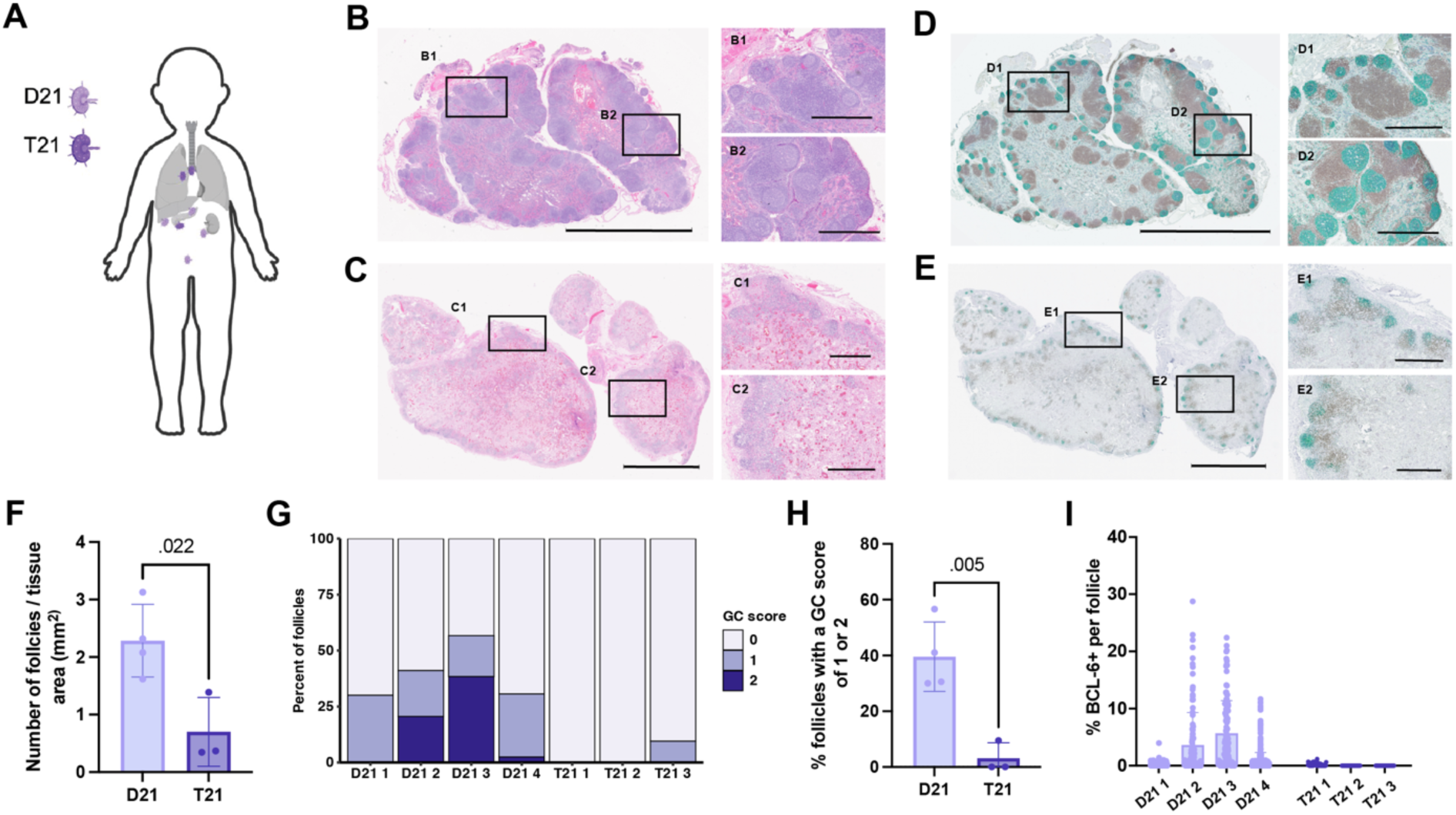
Phenotypic and immunohistochemistry analysis of D21 and T21 lymph nodes reveals a paucity of germinal center formation in T21 lymph nodes. **(A)** Schematic of lymph node sample location across the body, colored by D21 and T21 status. **(B)** Hematoxylin and eosin (H&E) staining of one D21 infant lymph node, D21 3. Scale bars, 5 mm on large image and 1 mm on small images (B1 – B2). **(C)** H&E staining of one T21 infant lymph node, T21 1. Scale bars, 5 mm on large image and 1 mm on small images (C1 – C2). **(D)** Immunohistochemistry (IHC) staining of one D21 infant lymph node, D21 3. Green CD20, Brown CD3, Purple BCL-6. Scale bars, 5 mm on large image and 1 mm on small images (D1 – D2). **(E)** IHC staining of one T21 infant lymph node, T21 1. Green CD20, Brown CD3, Purple BCL-6. Scale bars, 5 mm on large image and 1 mm on small images (E1 – E2). **(F)** Quantification of the number of follicles per lymph node, normalized to the size of the lymph node (mm^2^). Follicles were included in all calculations if more than 20% of cells within the follicle stained for CD20, as assessed via HALO. Significance was assessed using an unpaired, two-sided Welch’s t test. **(G)** Percentage of follicles with each GC score (0, 1, or 2), in each lymph node. Follicles received a score of 2 if they stained for BCL-6 and had a visible germinal center and mantle zone structure, a score of 1 if they either stained for BCL-6 or had a visible germinal center and mantle zone structure, and a score of 0 if they did not stain for BCL-6 nor had a visible germinal center and mantle zone structure. **(H)** Percentage of follicles per lymph node with a GC score of 1 or 2, split between D21 and T21 lymph nodes. Missing error bars indicate that the lower error bars extend below zero. Significance was assessed using an unpaired, two-sided Welch’s t test. **(I)** Percentage of BCL-6 positive cells per follicle, split by sample. Quantification conducted via HALO. Missing error bars indicate that the lower error bars extend below zero.

To corroborate these H&E observations, we conducted immunohistochemistry (IHC) staining to assess protein expression of CD20 (a marker of B cells), BCL-6 (a canonical marker of germinal center B cells^18^) and CD3 (a marker of T cells) (Figure 1D-E, S1D-E). IHC staining was quantified using HALO^TM^ image analysis software. Although there were no significant differences in CD3 staining between T21 and D21 lymph nodes (Figure S1F), there was a paucity, although not overt, of CD20 staining in T21 as compared to D21 lymph nodes (Figure S1G), perhaps attributable to the well-described deficit in the T21 B cell compartment in the peripheral blood.^11–15^ IHC confirmed the expected germinal center formation in D21 lymph nodes, with clear B cell follicles and T cell zones in all four lymph nodes (Figure 1D, S1D). In contrast, all three T21 lymph nodes had an almost complete lack of BCL-6 staining (Figure 1E, S1E). Quantification of B cell follicle numbers (including both primary and secondary follicles) demonstrated that T21 lymph nodes have significantly fewer follicles per mm^2^ (p-value = 0.022) (Figure 1F), at approximately half the follicle density of D21 lymph nodes. We then scored the primary and secondary follicles using both structural and staining information. Follicles received a score of 2 if they stained for BCL-6 and had a visible germinal center and mantle zone structure (secondary follicle), a score of 1 if they either stained for BCL-6 or had a visible germinal center and mantle zone structure (secondary follicle), and a score of 0 if they did not stain for BCL-6 nor had a visible germinal center and mantle zone structure (primary follicle) (Figure 1G). T21 lymph nodes had significantly fewer follicles that scored either a 1 or 2, with zero follicles that scored a 2 across all three lymph nodes (Figure 1G, H). Additionally, although D21 lymph nodes showed a spectrum of BCL-6 staining per follicle and between lymph nodes, as is expected with the formation of both primary and secondary follicles, all three T21 lymph nodes displayed an almost complete lack of BCL-6 staining in all follicles (Figure 1I). Combined, this H&E and IHC staining of T21 and D21 infant lymph nodes shows a striking paucity of germinal center formation in individuals with T21.

### Spatial transcriptomics confirms immune cell dysregulation in T21 lymph nodes

With this difference in mind, we conducted 10x Genomics Visium SD spatial transcriptomic analysis on six of these lymph nodes (n=3 D21 and n=3 T21) (Figure 2A) to further investigate morphological and structural differences between D21 and T21 lymph nodes. UMAP visualization revealed 10 clusters that aligned well with lymph node tissue regions, identifying the paracortex/T cell zones, B cell follicles, medulla (including the medullary sinus and medullary cords), subcapsular sinus, red blood cells/vascularity, and the capsule and surrounding muscle tissue (Figure 2A-C).

**Figure 2.**
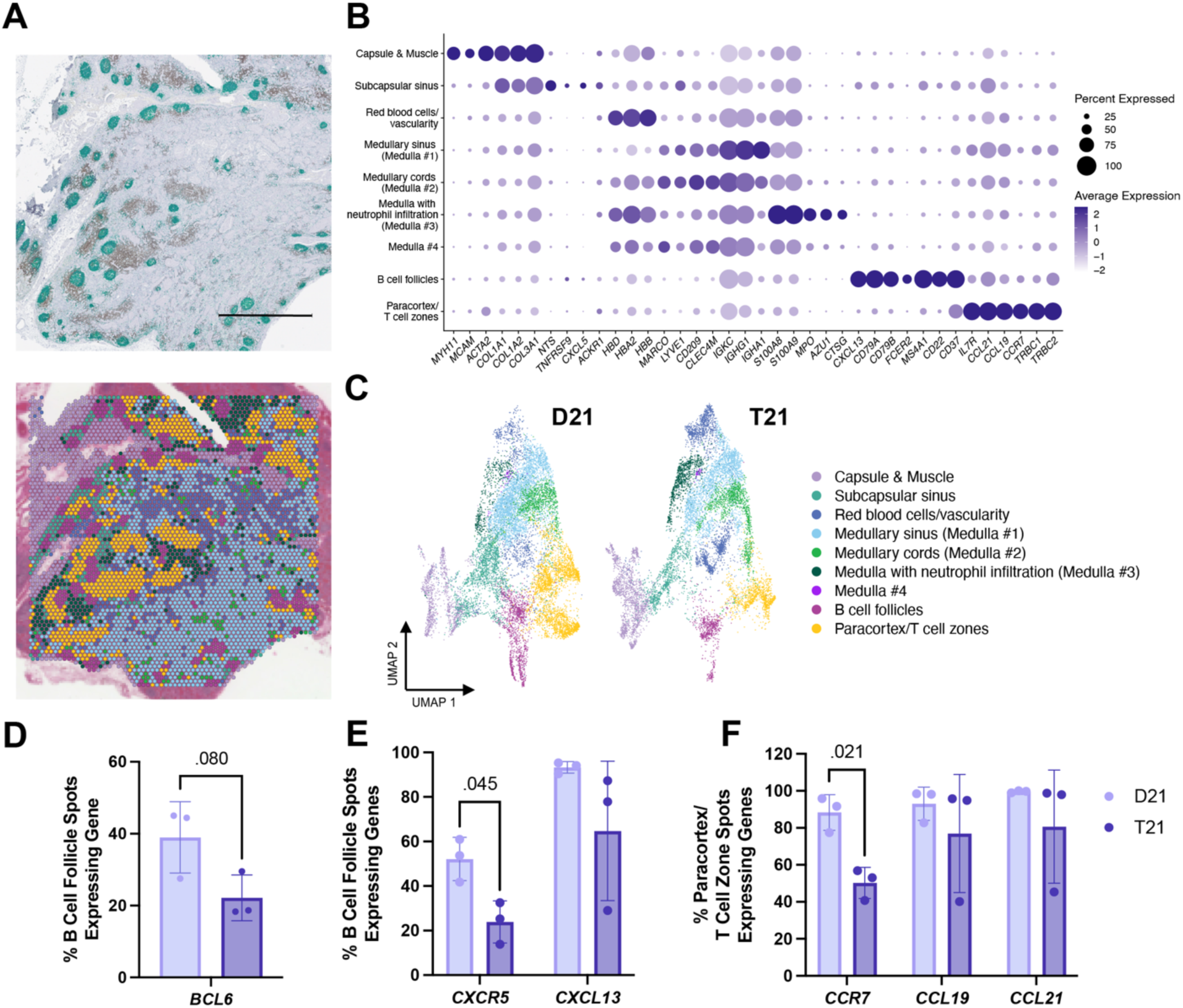
Spatial transcriptomics analysis of T21 and D21 lymph nodes. **(A)** Immunohistochemistry staining of D21 lymph node (Green CD20, Brown CD3, Purple BCL-6) and corresponding 10x Genomics Visium SD analysis from the same tissue block. Scale bar, 2 mm. **(B)** Dotplot of identifying markers for each cluster. Size of dot indicates the percent of spots expressing the gene, average expression indicated by color. **(C)** UMAP of clusters, labeled based on tissue histology and gene expression. **(D)** Barplot of the percentage of spots expressing *BCL6* per sample, within the B cell follicle cluster, compared between D21 and T21 samples. Significance was assessed using an unpaired, two-sided Welch’s t test. **(E)** Barplot of the percentage of spots expressing *CXCR5* and *CXCL13* per sample, within the B cell follicle cluster, compared between D21 and T21 samples. Significance was assessed using multiple unpaired, two-sided Welch’s t tests, adjusted using the Benjamini-Hochberg method. **(F)** Barplot of the percentage of spots expressing *CCR7, CCL19,* and *CCL21* per sample, within the paracortex/T cell zone cluster, compared between D21 and T21 samples. Significance was assessed using multiple unpaired, two-sided Welch’s t tests, adjusted using the Benjamini-Hochberg method.

To assign tissue regions to clusters, we used previously defined markers of human lymph node stromal cells and immune cells (Figure 2B, S2A). We identified a capsule and surrounding muscle tissue cluster (cluster 1), which showed high expression of muscle and collagen markers (*MYH11, MCAM, ACTA2*, *COL1A1, COL1A2, COL3A1*). We also identified a subcapsular sinus cluster (cluster 4), with high expression of collagen markers (*COL1A1, COL1A2, COL3A1)*, indicative of the conduit system of the lymph nodes,^20^ and markers of the ceiling and floor lymphatic endothelial cells found in the subcapsular sinus (*NTS, TNFRSF9, CXCL5, ACKR1*).^21^ The red blood cell/vascularity cluster (cluster 2) showed elevated expression of red blood cell genes, such as *HBD*, *HBA2*, and *HBB*. The medulla region consisted of 4 clusters, expressing markers of medullary sinus lymphatic endothelial cells (*LYVE1, CD209, MARCO, CLEC4M*^22^), as well as markers of B cells (*IGKC, IGHG1, IGHA1*), corresponding with the known presence of antibody-secreting plasma cells and B cells in the medullary cords (clusters 0, 5, 7 and 9). One medulla cluster (cluster 7) had particularly high expression of neutrophil markers (*S100A8, S100A9, MPO, AZU1, CTSG*), suggesting neutrophil infiltration. B cell follicles (cluster 6) were marked by expression of *CXCL13, CD79A, CD79B,* and other B cell markers, while two paracortex/T cell zones clusters (clusters 3 and 8) were distinguished via expression of *CCR7, CCL19, CCL21, IL7R, TRBC1,* and *TRCB2,* and were subsequently combined for future analysis (Figure 2B).

All clusters were present in both D21 and T21 samples, with no overt differences in cluster frequency between groups (Figure 2C, S2B-C). Spatial transcriptomics also confirmed the IHC findings: fewer B cell follicle spots expressed *BCL6* in the T21 lymph nodes compared to D21 lymph nodes (Figure 2D). Expression of *CXCR5,* a B cell migration receptor marker, was significantly decreased (p = 0.045) in the T21 B cell follicle spots, coupled with more variable expression of *CXCL13*, the corresponding migration chemokine, compared to the D21 B cell follicle spots (Figure 2E). This trend was mirrored in the T21 T cell zone spots: *CCR7,* a T cell migration receptor marker, was significantly decreased (p = 0.021), and expression of the corresponding chemokines, *CCL19* and *CCL21,* again showed much greater variability than in the D21 lymph nodes (Figure 2F). As these receptors and chemokines are important for migration within the lymph nodes, a decrease in expression within these regions may point towards a functional explanation for the paucity of germinal centers seen in these T21 lymph nodes.

### Spatial transcriptomics identifies stromal cell and follicular dendritic cell deficits in T21 lymph nodes

To further elucidate potential differences between the D21 and T21 lymph nodes, we sought to characterize the stromal architecture of the lymph node, including critical adhesion molecules ICAM1 and VCAM1, as well as specialized follicular dendritic cells (FDCs), lymphatic endothelial cells (LECs), and fibroblastic reticular cells (FRCs). First, as control markers, we assessed expression of *PAXBP1* and *BACH1*, genes on chromosome 21, and expression of *ATXN2* and *OTUB1*, two genes that are not on chromosome 21 and are associated with basic cellular assembly and function that should not differ significantly between groups. As anticipated, *PAXBP1* and *BACH1* showed around 1.5x expression in the T21 groups compared to the D21, while *ATXN2* and *OTUB1* showed similar expression between groups (Figure 3A).

**Figure 3.**
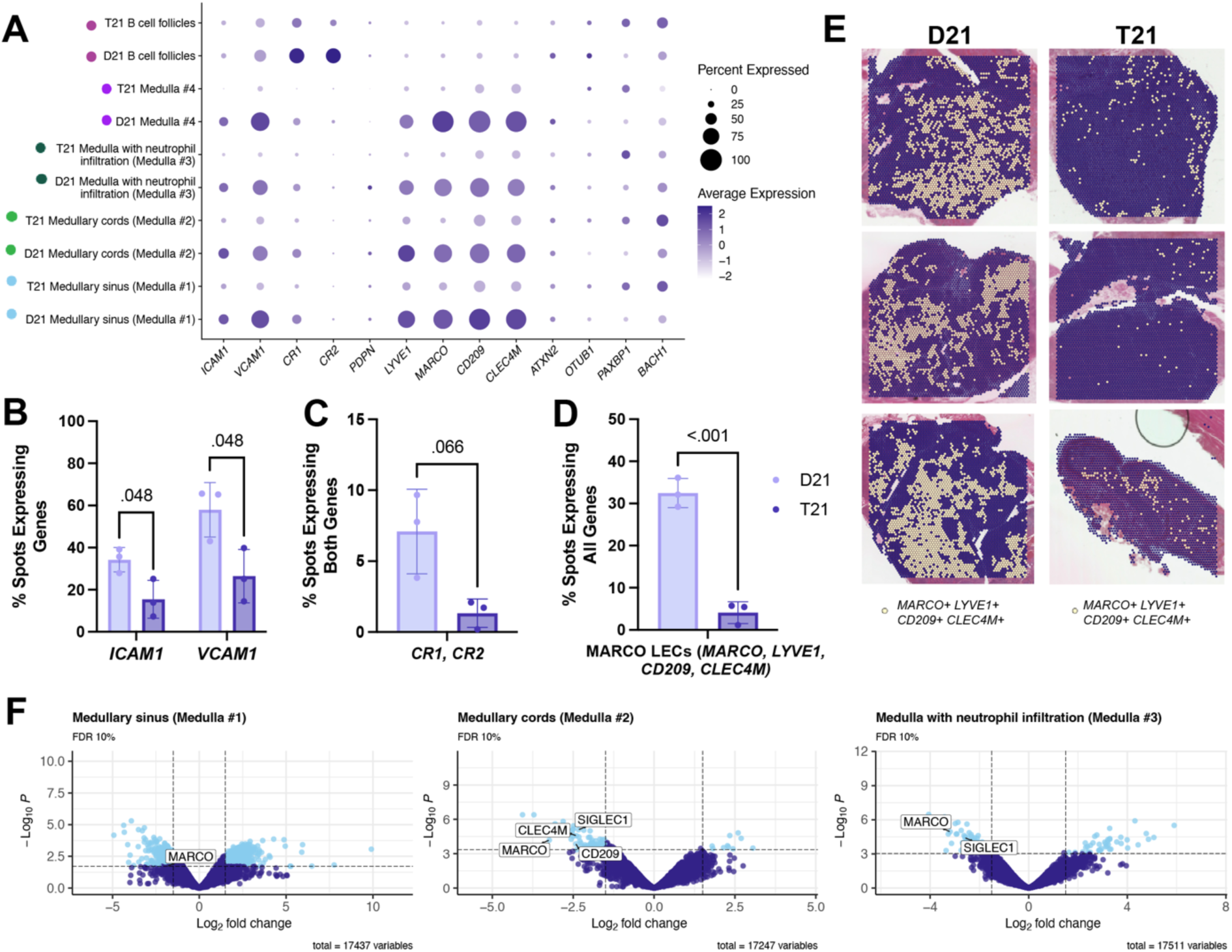
Spatial transcriptomics identifies stromal cell and follicular dendritic cell deficits in T21 lymph nodes. **(A)** Dotplot of stromal cell gene expression, split between medullary cell clusters and sample type (D21 and T21). Size of dot indicates the percent of spots expressing the gene, average expression indicated by color. **(B)** Barplot of the percentage of spots expressing *ICAM1* and *VCAM1,* per sample, compared between D21 and T21 samples. Significance was assessed using multiple unpaired, two-sided Welch’s t tests, adjusted using the Benjamini-Hochberg method. **(C)** Barplot of the percentage of spots expressing both *CR1* and *CR2,* per sample, compared between D21 and T21 samples. Significance was assessed using an unpaired, two-sided Welch’s t test. **(D)** Barplot of the percentage of spots expressing *MARCO, LYVE1, CD209,* and *CLEC4M* per sample, compared between D21 and T21 samples. Significance was assessed using an unpaired, two-sided Welch’s t test. **(E)** Visualization of spots expressing *MARCO*, *LYVE1*, *CD209*, and *CLEC4M*, per sample (as defined by gene expression > 0.5 for all markers). **(F)** Volcano plots for each medulla cluster. Significant DE genes are highlighted in light blue (FDR < 0.10, |log_2_ fold change| > 1.5). Medulla cluster #4 was excluded because it was not represented in all samples.

We identified striking differences in the expression of adhesion molecules between D21 and T21 lymph nodes. The average percentage of spots expressing *ICAM1* was decreased by 18.79% (p = 0.048) in T21 lymph nodes compared to D21 lymph nodes, while the average percentage of spots expressing *VCAM1* was decreased by 31.51% (p = 0.048), demonstrating a significant decrease in important cell adhesion molecules in T21 lymph nodes (Figure 3A, B). This reduction may explain the follicular hypocellularity and reduced follicular B cells observed in the T21 lymph nodes, as immune cells may be less prone to migrate properly into and within the lymph nodes. Next we evaluated FDCs, as they are essential for germinal center formation and drive antigen-specific B cell selection and proliferation.^23^ The average expression of *CR1* and *CR2*, markers of FDCs, was decreased in the B cell follicle cluster of T21 lymph nodes (Figure 3A), and the average percentage of spots expressing both markers was decreased in the T21 lymph nodes by 5.76% (p = 0.066), compared to D21 lymph nodes (Figure 3C). Given the essential role of FDCs in germinal center formation, the reduction of FDC markers in T21 lymph nodes is an important factor likely contributing to imperfect priming of adaptive immune responses and germinal center formation.

LECs and FRCs are important for maintaining lymph node structure, while also facilitating lymph flow, antigen exchange, and immune cell trafficking and positioning into and within the lymph node.^21,22^ To investigate stromal cell markers, we assessed expression of *LYVE1*, expressed on most LECs, and *PDPN*, a marker of FRCs.^21,24^ There were no notable differences in *PDPN* expression between D21 and T21 lymph nodes (Figure 3A, S3A), suggesting that some, but not all, stromal cells are affected in T21 lymph nodes. However, in all four medulla clusters (comprised of medullary sinus and medullary cords, as well broader medulla and medulla with neutrophil infiltration), T21 lymph nodes showed both decreased average expression and fewer spots expressing the LEC *LYVE1* marker (Figure 3A). Given this notable decrease in expression in the T21 medulla clusters, we further investigated this stromal subtype. LECs have been well-characterized in the human lymph node in recent years,^25,26^ revealing region-specific subsets with varying functions and distinguishing markers. We thus assessed expression of each known subset, examining ceiling and floor LECs (cLECs and fLECs), valve LECs (vLECs), PTX3-LECs in the paracortical sinuses, and MARCO-LECs in the medullary sinuses.^22^ Although gene expression of vLECs, fLECs, cLECs, and PTX3-LECs did not differ significantly between the medulla clusters of D21 and T21 lymph nodes (Figure S3B), we found a notable decrease in the expression of markers associated with MARCO-LECs (Figure 3A).

MARCO-LECs are found in the medullary sinuses and are important for virus scavenging, antigen archiving, and supporting the medullary sinus macrophages.^22^ They can be identified via expression of *MARCO* and *LYVE1*, as well as high levels of *CD209* and *CLEC4M*. In all 4 medulla clusters, we saw decreases in expression of *MARCO*, *LYVE1*, *CD209*, and *CLEC4M*, as well as a significant decrease in the number of spots expressing all four markers together (Figure 3A, D, E). In addition to assessing expression at the single spot level, we conducted pseudobulk analysis of each spatial cluster to more rigorously assess differential expression. We observed a significant decrease in the expression of markers identifying MARCO-LECs, including *MARCO*, *CD209*, and *CLEC4M,* in the medullary sinus (medulla #1), medullary cords (medulla #2), and medulla with neutrophil infiltration clusters (medulla #3) (FDR < 0.10, |log_2_FC| > 1.5) (Figure 3F). Medulla cluster #4 was excluded from analysis, as it did not appear in all samples. Additionally, given the role that MARCO-LECs play in supporting lymph node macrophages, we assessed expression of *SIGLEC1,* a macrophage marker expressed by both subcapsular sinus macrophages (SCM) and medullary sinus macrophages (MSM). *SIGLEC1* expression was also significantly decreased in medulla cluster #2 and #3 (Figure 3F). In combination, this suggests that a reduction of MARCO-LECs, SCMs, and MSMs may further exacerbate the architectural dysfunction observed in T21 LNs.

Taken together, decreased expression of critical cellular adhesion molecules, fewer FDCs, and a striking lack of MARCO-LECs in T21 lymph nodes points towards a broad deficit in stromal support. Stromal cells in the lymph nodes play a critical role in recruiting cells to their appropriate site, antigen exchange, and immune cell function, and a stromal cell deficit may contribute to the paucity of germinal centers seen in T21 lymph nodes. In particular, follicular dendritic cells are known to support germinal center responses through antigen retention and presentation to B cells — FDCs are important for extended antigen presentation, allowing B cells to undergo affinity maturation. FDCs also promote B cell survival in the germinal centers.^23^ Furthermore, lymph node macrophages play a role in antigen processing and presentation; complement-opsonized antigen is bound by SCMs and is transferred to the FDCs. It has also been shown that antigen-specific B cells can bind their antigen directly on SCMs, triggering antigen internalization, presentation to T cells, and germinal center formation.^27^ If these macrophages are not properly supported by MARCO-LECs or are fewer in number, this may further contribute to the decrease in germinal center formation found in the T21 lymph nodes.

### Single cell gene expression analysis of T21 B cells reveals an activated memory compartment

Given the reduced germinal center formation seen in T21 lymph nodes and deficits in markers of supportive immune and stromal cells, we next sought to profile the peripheral B cell compartment in unrelated individuals with T21 via paired single-cell RNA-sequencing (RNA-seq) and V(D)J sequencing (BCR-seq) to further investigate B cell differences in T21. Sorted CD19+ B cells from six T21 adults and four age- and sex-matched D21 adults (Figure 4A, S4A) were assayed, allowing us to capture 90,782 high-quality single-cell transcriptomes (n = 44,230 cells from D21 individuals and n = 46,552 cells from T21 individuals).

**Figure 4.**
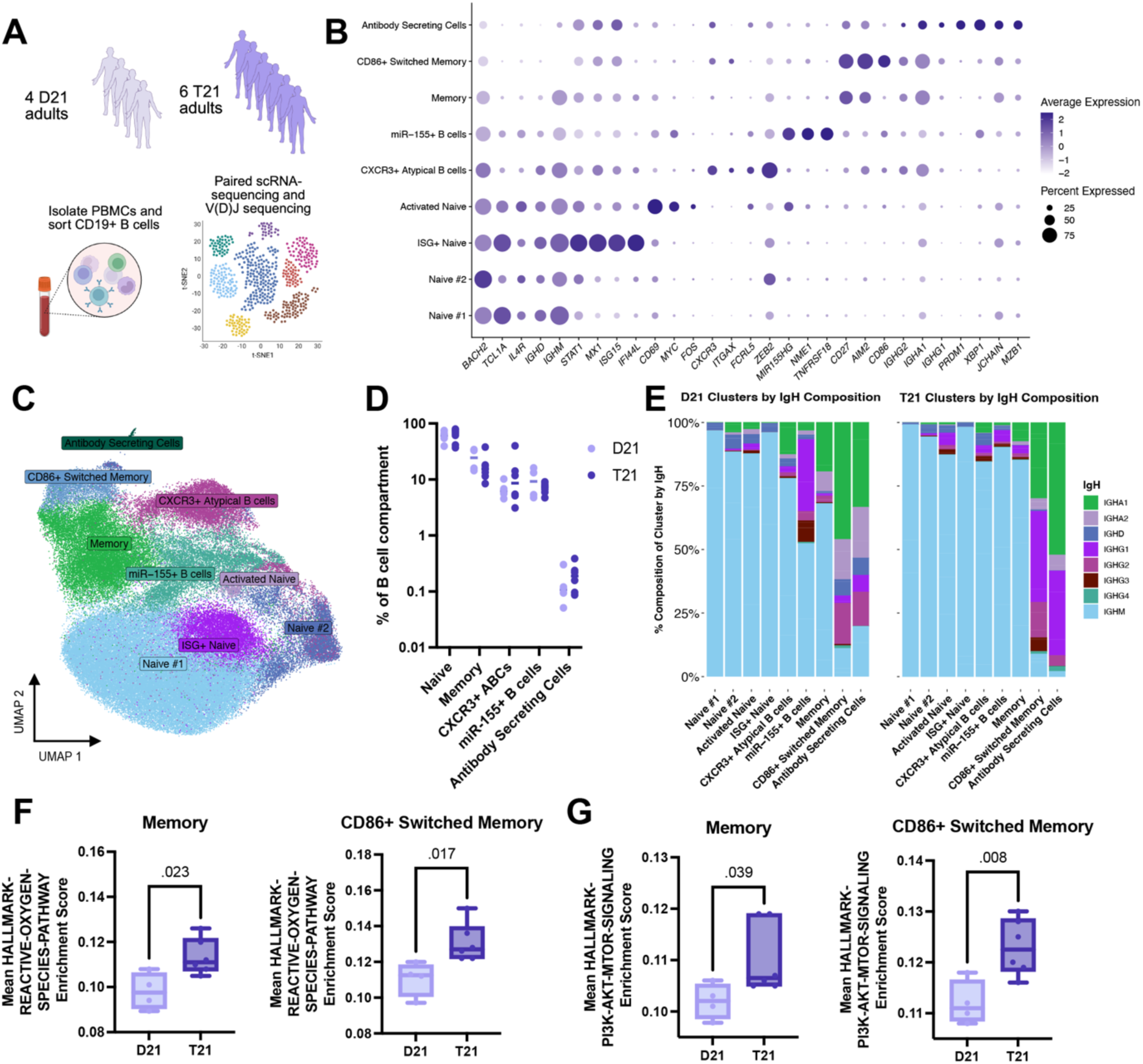
T21 B cells upregulate pathways of activation and apoptosis compared to D21 B cells. **(A)** Diagram of experimental setup. **(B)** Dotplot of identifying markers for each cluster. Size of dot indicates the percent of spots expressing the gene, average expression indicated by color. **(C)** UMAP of all samples, labeled via cluster. **(D)** Each cluster (naïve and memory clusters are summed together), as a percentage of the total B cell compartment, split between D21 and T21 samples. Log10 scale. **(E)** Distribution of IGH class per cluster, split between D21 and T21 samples. **(F)** Mean Hallmark Reactive Oxygen Species pathway gene set enrichment score calculated from memory B cells and CD86+ switched memory B cells, split between D21 and T21 samples. Significance was assessed using an unpaired, two-sided Welch’s t test. **(G)** Mean Hallmark PI3K-AKT-MTOR Signaling gene set enrichment score calculated from memory B cells and CD86+ switched memory B cells, split between D21 and T21 samples. Significance was assessed using an unpaired, two-sided Welch’s t test.

Normalization, Harmony integration,^28^ and clustering yielded 10 B cell clusters (Figure 4B-C). Two clusters were composed of naïve B cells (Naïve #1 and Naïve #2), expressing markers including *BACH2, TCL1A,* and *IL4R*^29–32^; additionally, we identified a type I interferon stimulated gene (ISG+) naïve cluster (expressing *STAT1, ISG15, MX1,* and *IFI44L*^32^) and an activated naïve cluster (expressing *CD69* and *MYC*^32^). Two clusters were defined by *ZEB2, ITGAX,* and *FCRL5,*^31–33^ markers associated with a B cell compartment known as atypical B cells (ABCs). We have previously reported CD11c+ ABCs to be increased in individuals with T21,^12^ while others have found this population in the context of chronic infection and autoimmunity.^34^ The second ABC-like cluster, in addition to *ZEB2, ITGAX,* and *FCRL5* expression, also displayed expression of *CXCR3,* which has been reported as upregulated in ABCs across clinical phenotypes.^12,33,34^ In subsequent analyses, these two clusters were combined into a single CXCR3*+* atypical B cell cluster (Figure 4B-C). Because cells expressing high levels of *ZEB2, ITGAX,* and *FCRL5* were distributed across clusters, we computed a gene set score for all cells and assigned cells to the CXCR3*+* atypical B cell cluster if they scored above a set threshold (see Methods) (Figure 4B-C).

We identified a B cell population characterized by high expression of *MIR155HG*, *NME1*, and *TNFRSF18* (miR-155+ B cell cluster). A similar population has been reported previously in human tonsillar B cells.^35,36^ *MIR155HG* and *NME1* are both known to play a role in class-switching,^37,38^ suggesting these cells are likely in the process of class switching. Furthermore, we identified two clusters of memory B cells (Memory and CD86+ Switched Memory), both expressing *CD27* and *AIM2,* as well as markers of class switched B cells, such as *IGHA1* and *IGHG2*, with the CD86+ Switched Memory cluster expressing higher levels of *CD86* and much higher usage of switched Ig heavy chains (Figure S4B).^29,30,32^ Finally, we identified a small cluster of antibody-secreting cells, defined by expression of *PRDM1*, *XBP1*, *JCHAIN*, and *MZB1*^30^ (Figure 4B-C). All together, these clusters reflect the known cell type composition of the B cell compartment in PBMCs; 60-70% of the samples are identified as naïve B cells and around 10-30% as memory, with smaller CXCR3+ atypical B cell, miR-155+ B cell, and antibody secreting cell compartments (Figure 4D).

Next, we directly compared D21 and T21 B cells. Distribution of the B cell compartments did not differ overtly between D21 and T21 samples (Figure 4D, S4C), although there was a slight expansion in naïve B cells and reduction in the memory compartment, as expected in T21 individuals.^39^ Interestingly, the T21 samples on average displayed a higher percentage of cells whose VDJ region mapped to the IGHM heavy chain and fewer cells that mapped to the class-switched regions (IGHA1, IGHA2, IGHG2, IGHG3, IGHG4, with IGHG1 being the singular exception) (Figure 4E, S4D). This observation aligns with clinical information known about individuals with T21, as they have been shown to exhibit fewer class-switched memory B cells and a less potent and durable B cell response.^39^

Functionally, T21 Memory and CD86+ Switched Memory B cell clusters showed significantly increased UCell^40^ enrichment scores for the reactive oxygen species (ROS) and PI3K-AKT-MTOR signaling pathways as compared to D21 (Figure 4F, G). Interestingly, ROS signaling is critical in the BCR signaling pathway: ROS expression is higher in activated B cells and ROS production drives B cell apoptosis, presenting a possible mechanism for the significantly smaller B cell compartment in T21 individuals.^41^ Furthermore, high PI3K/AKT expression is associated with lower FOXO expression, favoring plasmablast differentiation.^42^ Individuals with T21 are known to have a significantly expanded plasmablast compartment suggestive of autoimmunity, further confirming the clinical relevance of this increased pathway signature.^12^ Additionally, patients with mutations in *PIK3CD or PIK3R1,* known as activated PI3K delta syndrome (APDS), exhibit clinical parallels to T21, with respiratory tract infections, autoimmune disorders, and developmental delays.^43^ Therefore, this elevated PI3K-AKT-MTOR pathway may then be important in the biological and clinical outcomes in T21.

### BCR sequencing of T21 B cell compartment reveals critical changes in somatic hypermutation and autoimmune phenotypes

To further understand outcomes of B cell education in individuals with T21, we assessed somatic hypermutation frequency of the immunoglobulin heavy chain variable region (IGHV) using BCR-sequencing. We first confirmed that somatic hypermutation rates increased from naïve to memory compartments and found the highest levels of somatic hypermutation in the CD86+ switched memory B cell compartment (Figure S5A), as is biologically expected. Strikingly, we found a decrease in somatic hypermutation levels of the memory compartment in T21 samples compared to D21 samples (p = 0.051) (Figure 5A, B). This decrease in somatic hypermutation was maintained after pooling together memory and CD86+ switched memory cells (Figure 5C).This finding is indeed suggestive of a deficit in B cell education in individuals with T21, which we first noted above in infant lymph nodes (Figure 1) and that has been clinically observed via dysregulation in the response to vaccination that is seen in individuals with T21.^7–10^ This suggests that adaptive immune responses against infectious disease develops in T21 but may not be as mature as compared to D21 individuals.

**Figure 5.**
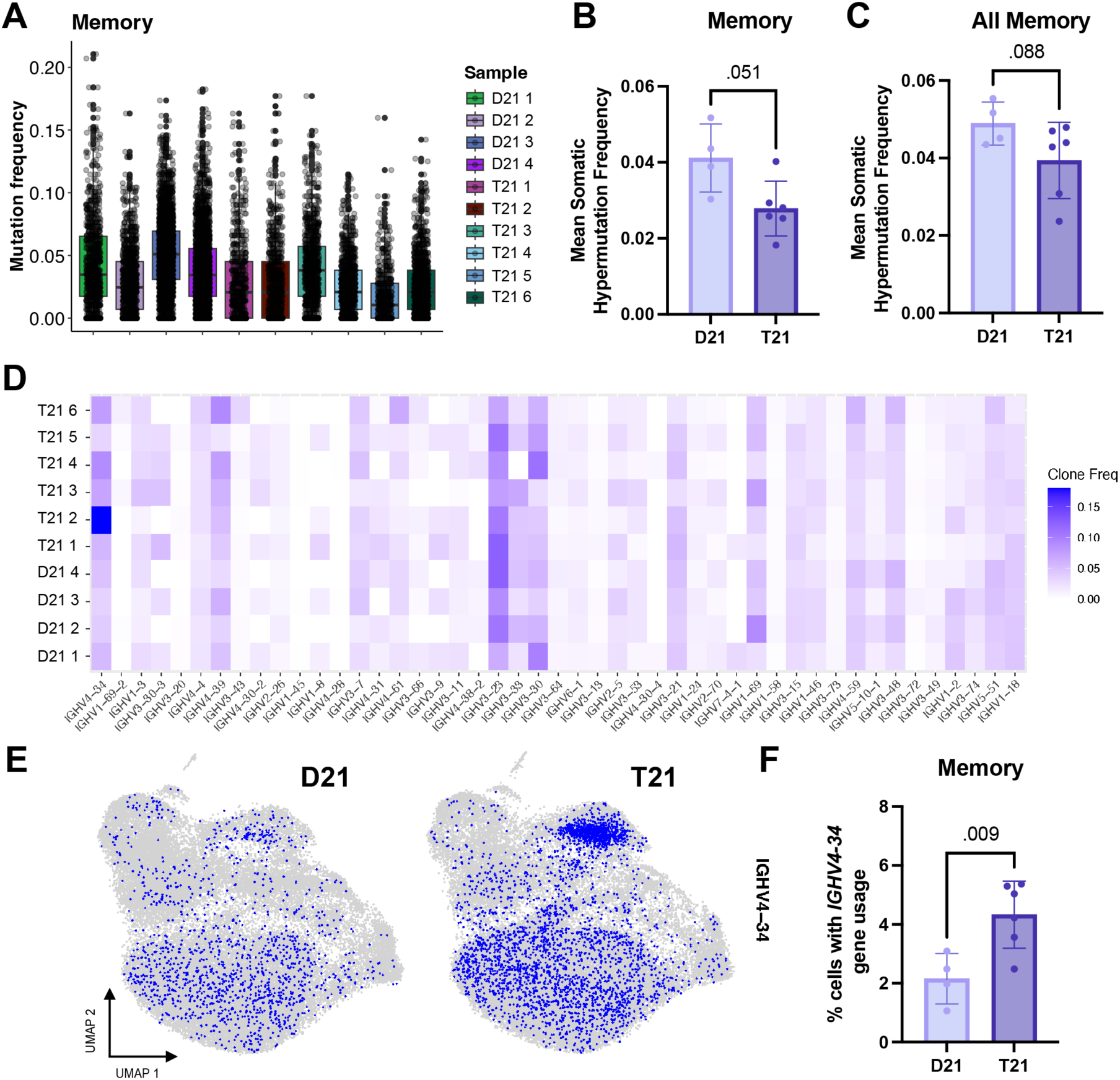
BCR sequencing of T21 B cell compartment reveals critical changes in somatic hypermutation and autoimmune phenotypes. **(A)** Somatic hypermutation frequency of immunoglobulin heavy chain variable region (IGHV) from memory B cells, separated into each sample. **(B)** Mean somatic hypermutation frequency of memory B cells per sample, split between D21 and T21. Significance was assessed using an unpaired, two-sided Welch’s t test. **(C)** Mean somatic hypermutation frequency of combined memory B cells and CD86+ switched memory B cells per sample, split between D21 and T21. Significance was assessed using an unpaired, two-sided Welch’s t test. **(D)** Heatmap of variable (V) gene usage, per sample. Clonal frequency of each V gene is indicated by color. **(E)** UMAP of cells with *IGHV4-34* gene usage (indicated in blue), split between D21 and T21 samples. **(F)** Percentage of memory B cells aligned to the *IGHV4-34* heavy chain gene per sample, split between D21 and T21. Significance was assessed using an unpaired, two-sided Welch’s t test.

Conversely, individuals with T21 exhibit an increased prevalence of autoimmunity, which we and other have noted.^12,39^ We thus assessed differential usage of the heavy chain V gene in our cohort (Figure 5D). There was a notable increase in usage of the *IGHV4-34* heavy chain gene in the T21 samples, which has a known autoimmune function and plays a critical role in systemic lupus erythematosus (Figure 5E).^44^ In particular, the memory B cell compartment had significantly increased usage of the *IGHV4-34* heavy chain gene (p = 0.009) (Figure 5F). This expansion of a known autoimmune B cell population also aligns with T21 clinical data, as these individuals have a significantly increased autoimmune disease burden. Ultimately, assessment of the B cell compartment in individuals with T21 provides likely mechanisms for B cell dysfunction, including dysregulation in the rate of somatic hypermutation and the expansion of a putatively autoreactive B cell population.

## Discussion

Despite improved clinical care, individuals with T21 still experience a disproportionate burden of infectious disease and autoimmunity compared to typical individuals, with influenza infections and pneumonia being the third leading cause of death.^45^ In considering how to further extend lifespans and improve quality of life in the Down syndrome community, furthering our knowledge of the immune system and how it functions is critical to informing effective clinical care and treatment.

Herein we document for the first time structure and function of lymph nodes in T21 individuals. While typical infants should form germinal centers upon first exposure to microbes and vaccines in early life, experimentally this was shown in lung-associated lymph nodes as early as 6 months.^46^ In contrast, we document a paucity of germinal center formation in the T21 lymph nodes we examined here. In addition, spatial transcriptomics revealed a striking decrease in markers of adhesion molecules, follicular dendritic cells, and MARCO-lymphatic endothelial cells in the T21 lymph nodes. While D21 lymph nodes have the necessary stromal cells to support germinal center formation and B cell education, the T21 lymph nodes do not appear to, revealing a broader dysregulation of the lymph node structure, stromal support, and immune cell function.

The T21 lymph nodes we examined were derived from autopsies and while none have immune related causes of death (Supplemental Figure 1A), it is possible that our findings fall on the extreme end of the phenotypic spectrum of germinal center formation and B cell education in the T21 population. As more lymph nodes are examined across the lifespan in T21, it is likely that we will discover that individuals with T21 have a significantly increased variance in the numbers of germinal centers formed and possibly a delay in the ability to form germinal centers, as opposed to presenting with a complete germinal center formation deficiency. Significant clinical variability in T21 is a norm, as we and others have documented a wide spread of cytokinopathy, autoimmunity, and peripheral blood cell dysregulation ranging from typical-like to severe. It is of value to expand the investigation of T21 lymph nodes across age and clinical severity of autoimmunity and immune deficiency. Despite this, we anticipate that our samples likely reflect a broader germinal center dysfunction in individuals with T21.

To complement this in-depth analysis into an infant cohort, we expanded our investigation to a population of unrelated adults with T21. There, we examined sorted CD19+ B cells from PBMCs and performed paired single-cell RNA sequencing and V(D)J sequencing. We indeed confirm a dysregulated B cell compartment, with decreased somatic hypermutation rates and an increase in autoreactive *IGHV4-34* heavy chain usage. Additionally, memory B cells have elevated expression of pathways associated with activation and apoptosis, suggesting a further functional dysregulation, likely resulting from intrinsic and other peripheral immune defects. These results mirror our clinical understanding of T21, as B cells are still undergoing some version of specialization, but at a rate lower than that seen in typical individuals, and with dysregulation that may have to do with extrafollicular education resulting in autoimmunity.

Ultimately, our data advances understanding of B cell education in individuals with trisomy 21 in infancy and in adulthood. Given the paucity of germinal center formation, coupled with lower somatic hypermutation rates, and clinical data suggesting some amount of variability in potency and duration of vaccine response, it may be appropriate to consider clinical trials to assess an increased immunization schedule in individuals with T21. Increased vaccination may help protect against infectious disease and allow the immune system in individuals with T21 more opportunity to mount an effective protective response. Additionally, larger studies across age and clinical severity are needed to elucidate the spread of B cell education across the T21 cohort. Understanding how the immune response in T21 individuals stratifies across variables, such as immune population dysregulation, response to specific foreign antigen, and autoimmune presence, may allow us to more precisely understand the variability of the T21 immune system and best address the clinical needs of the T21 community.

## Data Availability

All raw sequencing data and histological data produced in the present study will be available upon formal journal publication.

## Acknowledgements

We thank all study participants for their contributions. We thank Dr. Monika Garcia-Barros and the team at the pathology department of the Icahn School of Medicine at Mount Sinai for their help with histology and immunohistochemistry staining. We thank Dr. Erin Bush and the team at the Columbia Genome Center of Columbia University Irving Medical Center for their help with the 10x Genomics paired single-cell RNA sequencing and V(D)J sequencing. This research was funded in part through the NIH/NCI Cancer Center Support Grant P30CA013696 and used the Genomics and High Throughput Screening Shared Resource. This publication was supported by the National Center for Advancing Translational Sciences, National Institutes of Health, through Grant Number UL1TR001873. We thank Juan Idiarte and the staff of the Columbia Stem Cell Initiative Flow Cytometry Core Facility, under the leadership of Michael Kissner, at Columbia University Irving Medical Center for their contributions to the work presented in this manuscript. This work was completed in part with Minerva computational resources provided by Scientific Computing and Data at the Icahn School of Medicine at Mount Sinai (Clinical and Translational Science Awards [CTSA] grant UL1TR004419, National Center for Advancing Translational Sciences). This work was also completed in part with computational resources provided by the Columbia University Center for Computational Biology and Bioinformatics (C2B2). Figures 1A and 4A were created with BioRender. The content is solely the responsibility of the authors and does not necessarily represent the official views of the NIH.

## Funding

R01AI150300, R01AI127372, P01AI186771, R01AI167802.

## Author contributions

D.B. supervised the study. M.R., O.S., A.A., F.K., and D.B. designed the experiments. S.B. coordinated the Down syndrome cohort and processed whole-blood samples. A.K. and R.B. helped acquire pathology samples. M.R. performed HALO analysis. M.R. and H.E.R. performed computational analyses. G.H.L. performed spatial transcriptomics sample preparation and sequencing. M.R. and D.B. wrote the manuscript, with input from all authors.

## Disclosures/competing interests

D.B. reports ownership in Lab11 Therapeutics.

---

**Figure S1.**
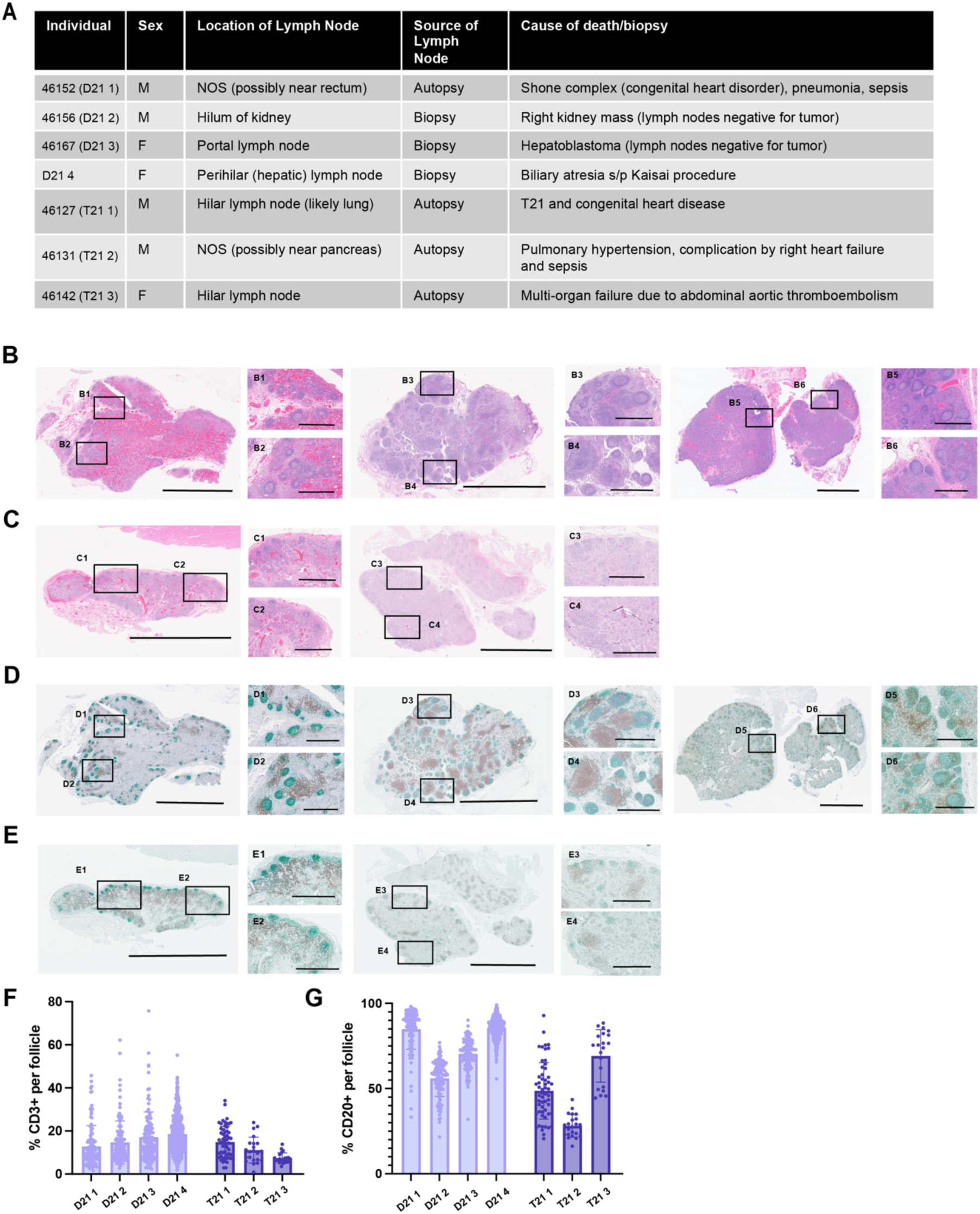
Phenotypic and immunohistochemistry analysis of D21 and T21 lymph nodes reveals a lack of germinal center formation in T21 lymph nodes. **(A)** Table containing lymph node label, sex, location in the body, and clinical phenotype of each donor. All samples are within 0-4 years old. **(B)** Hematoxylin and eosin (H&E) staining of D21 infant lymph nodes, D21 1 (B1-B2), D21 2 (B3-B4), and D21 4 (B5-B6). Scale bars, 5 mm on large images and 1 mm on small images (B1 – B6). **(C)** H&E staining of T21 infant lymph nodes, T21 3 (C1-C2) and T21 2 (C3-C4). Scale bars, 5 mm on large images and 1 mm on small images (C1 – C4). **(D)** Immunohistochemistry (IHC) staining of D21 infant lymph nodes, D21 1 (D1-D2), D21 2 (D3-D4), and D21 4 (D5-D6). Green CD20, Brown CD3, Purple BCL-6. Scale bars, 5 mm on large images and 1 mm on small images (D1 – D6). **(E)** IHC staining of T21 infant lymph nodes, T21 3 (E1-E2) and T21 2 (E3-E4). Green CD20, Brown CD3, Purple BCL-6. Scale bars, 5 mm on large images and 1 mm on small images (E1 – E4). **(F)** Percentage of CD3 positive cells per follicle, split by sample. Quantification conducted via HALO. **(G)** Percentage of CD20 positive cells per follicle, split by sample. Quantification conducted via HALO.

**Figure S2.**
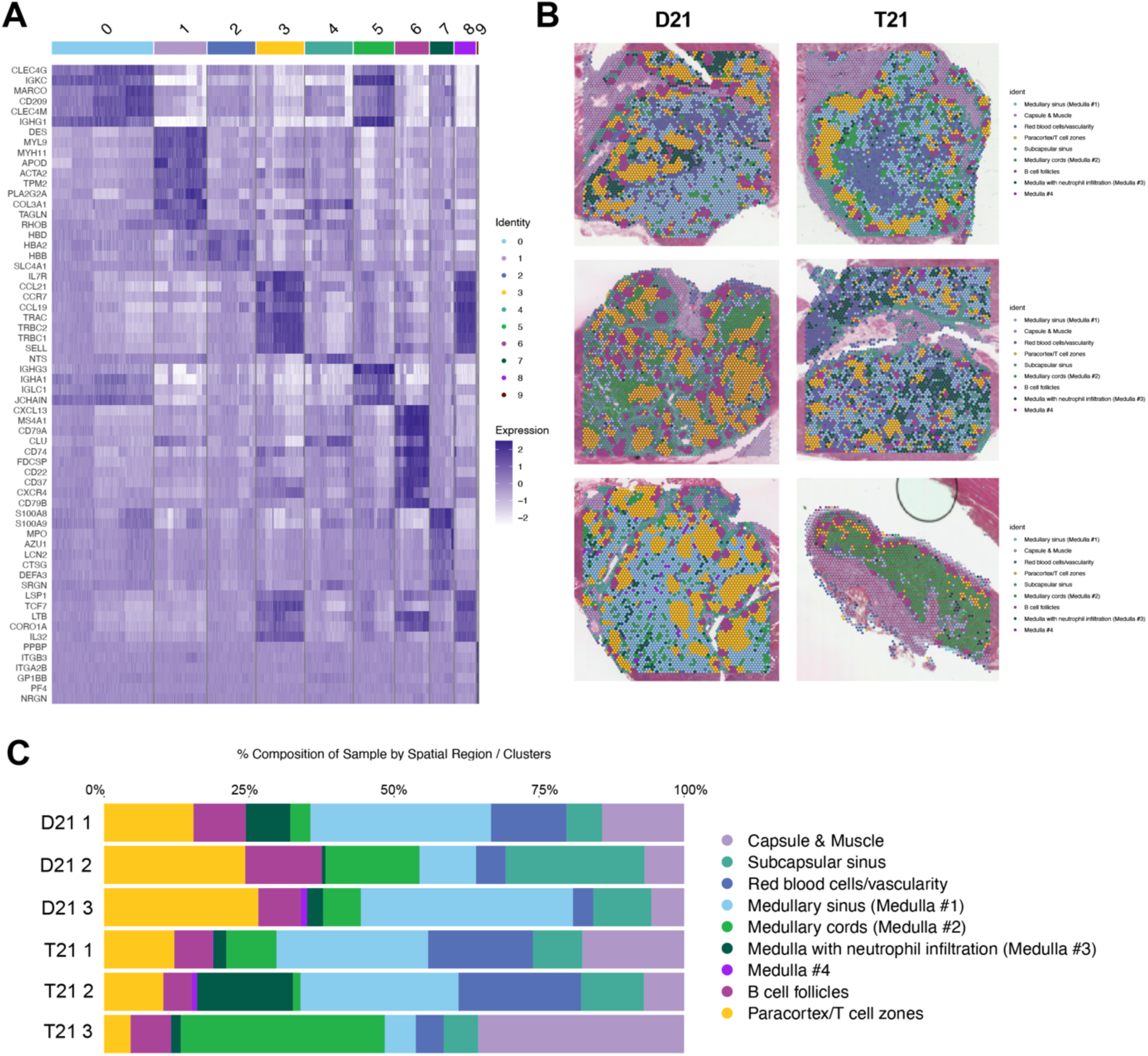
Spatial transcriptomics analysis of T21 and D21 lymph nodes. **(A)** Heatmap of top expressed genes in each UMAP cluster, prior to cluster labeling. **(B)** Visualization of spot assignment, per sample. **(C)** Percent composition of each sample by spatial cluster.

**Figure S3.**
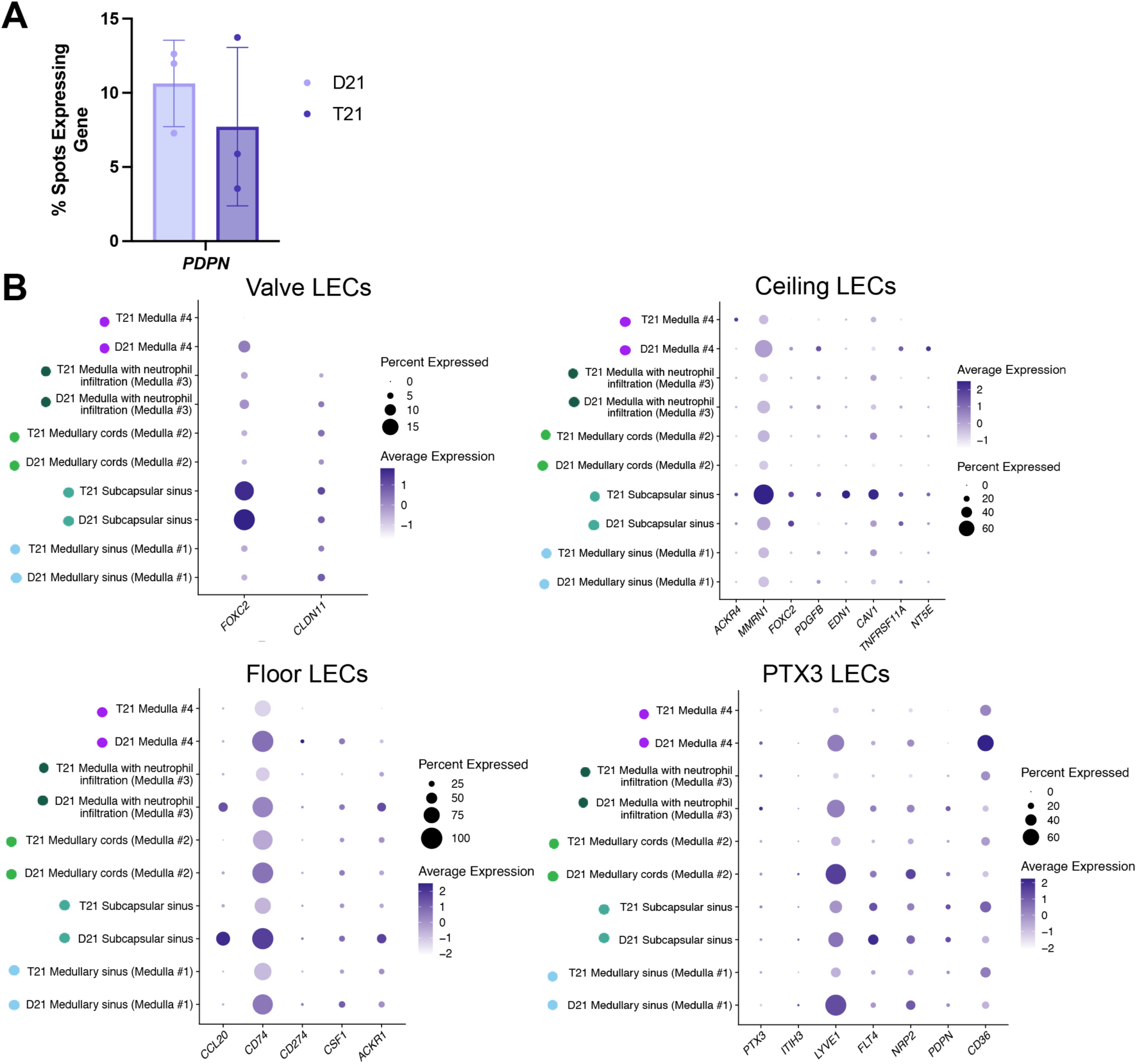
Stromal cell analysis. **(A)** Barplot of the percentage of spots expressing *PDPN* per sample, compared between D21 and T21 samples. **(B)** Dotplot of lymphatic endothelial cell (LEC) subtype (valve, ceiling, floor, and PTX3+) gene expression, split between stromal cell clusters and sample type (D21 and T21). Size of dot indicates the percent of spots expressing the gene, average expression indicated by color.

**Figure S4.**
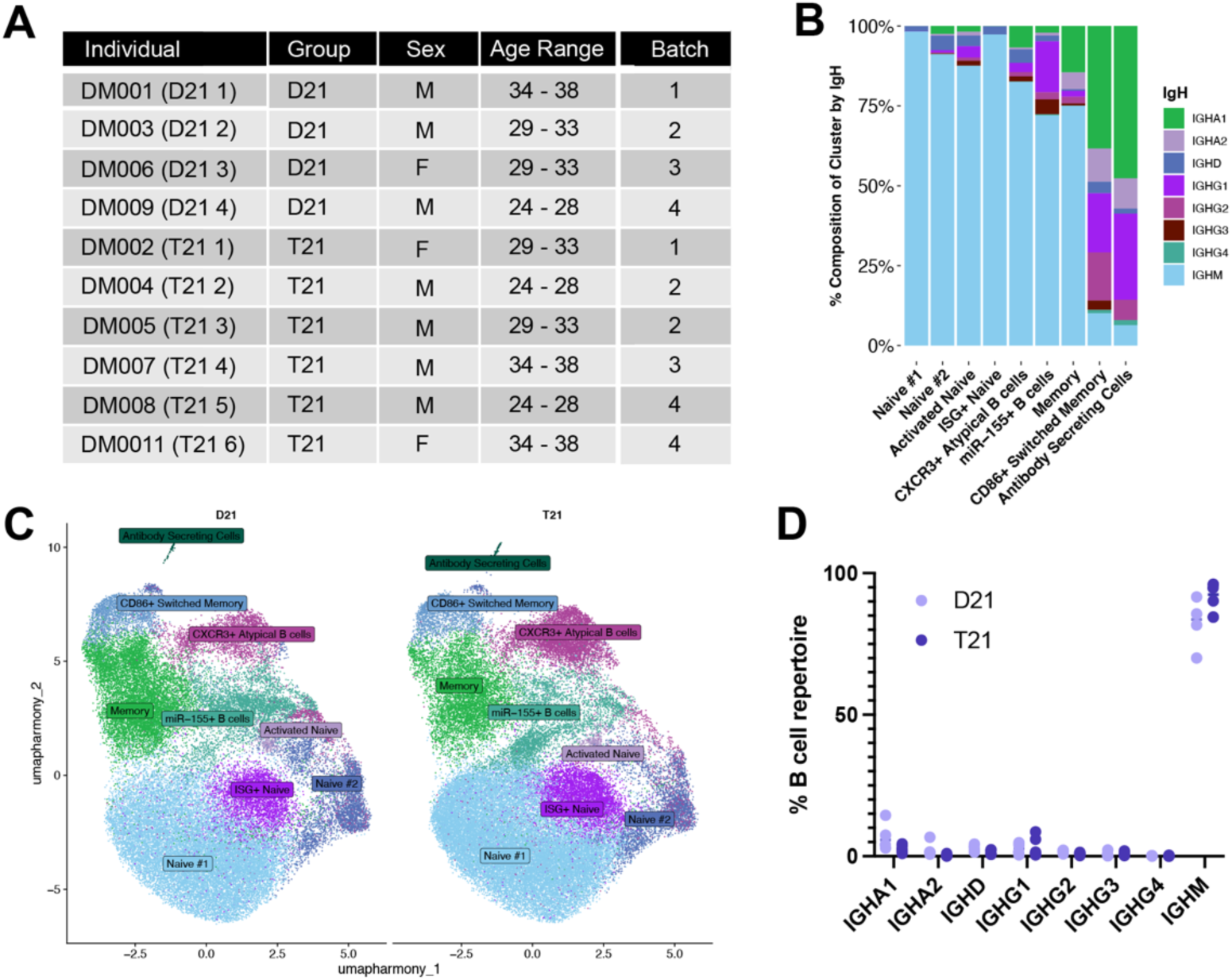
scRNAseq and BCRseq analysis of D21 and T21 B cells. **(A)** Label, group (D21 or T21), sex, and age range of all donors used for scRNAseq and paired V(D)J sequencing. **(B)** Distribution of IGH class per cluster. **(C)** UMAP of all samples, labeled via cluster, split between D21 and T21 samples. **(D)** Percentage of B cell repertoire mapping to each IgH gene, split between D21 and T21 samples.

**Figure S5.**
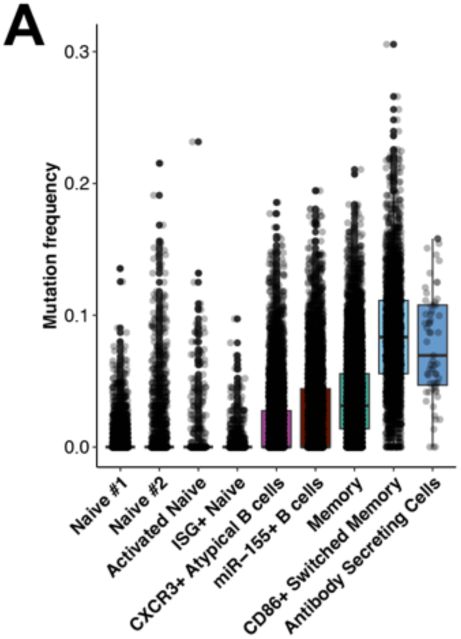
BCRseq analysis of D21 and T21 samples. **(A)** Somatic hypermutation frequency of immunoglobulin heavy chain variable region (IGHV) per cell, split by gene expression cluster.

## Materials and Methods

### PBMC acquisition

The study was approved under protocols at Mount Sinai Health System (MSHS) (IRB-18-00638/ STUDY-18-00627) and Columbia University Medical Center (CUIMC) (IRB-AAAV1815). All individual and sample IDs are de-identified and unknown to those outside of the research group. Patients were approached in person at the hospital or by email obtained through the NIH’s DS-Connect national registry (https://dsconnect.nih.gov). All patient data were deidentified. Written informed consent for all individuals in this study was provided in compliance with an institutional review board protocol. Control (D21) samples were obtained from the New York Blood Center. From each donor, blood was drawn into a cell preparation tube with sodium heparin (BD Vacutainer). Peripheral blood mononuclear cells (PBMCs) and plasma were isolated via Histopaque-1077 separation (Sigma-Aldrich, cat no. H8889) and subsequently stored in liquid nitrogen and at -80C, respectively, until use.

### Lymph node acquisition

Tissue samples were acquired from the Mount Sinai Pathology Department and Columbia University Biobank (IRB-AAAV1815). All patient data was deidentified and samples anonymized prior to receipt of the specimens. Lymph nodes were residual specimens from clinical care derived from autopsies and biopsies; sex, location of lymph node, and cause of death/biopsy are listed in Supplemental Figure 1A. Sample IDs listed in Supplemental Figure 1A are not known to anyone outside of the research group.

## Tissue Immunopathology

Formalin-fixed paraffin-embedded lymph nodes were cut into 5 μm sections and stained by the Biorepository and Pathology Core (ISMMS).

### Hematoxylin and eosin staining

Tissues were stained with hematoxylin and eosin to visualize nuclei and tissue structure.

### Multiplex immunohistochemistry staining

For sequential chromogenic multiplex IHC, the following markers were evaluated (CONFIRM anti-CD3 (2GV6) Rabbit Monoclonal Primary Antibody Cat. # 790-4341, CONFIRM anti-CD20 (L26) Primary Antibody Cat. # 760-2531, and bcl-6 (GI191E/A8) Mouse Monoclonal Antibody Cat. # 760-4241), prediluted from Roche using VENTANA DISCOVERY ULTRA automated slide staining instrument (Roche, Basilea, Switzerland). This system allows for automated baking, deparaffinization and cell conditioning. Each primary marker was followed by the correspondent secondary antibody (DISCOVERY Anti-Rabbit HQ Catalog # 760-4815, DISCOVERY OmniMap anti-Rb HRP (RUO) Catalog # 760-4311, DISCOVERY Anti-Mouse HQ Catalog # 760-4814) and the signal was developed with a different color: DISCOVERY ChromoMap DAB kit (RUO) Catalog # 760-159 (brown), DISCOVERY green HRP Kit Catalog # 760-271 (green), and DISCOVERY Purple kit (RUO) Catalog # 760-229 (purple). Importantly, after each sequential staining, slides went throughout a step of inhibition, heat denaturation, and neutralization. Tissues were counterstained with Hematoxylin to visualize the nuclei. Whole tissue sections on the slide were converted into high-resolution digital data using a NanoZoomer S210 Digital slide scanner (Hamamatsu).

### Immunohistochemistry quantitative analysis

The HALO^TM^ image analysis platform was used for quantitative tissue analysis (Indica Labs, Inc). The multiplex IHC module, in conjunction with the nuclear segmentation module, were used to segment nuclei and separate chromogenic stains to set up the system for quantitative analysis.

Follicles were selected on each lymph node and the multiplex IHC module was applied to assess staining for CD3, CD20, and BCL-6. Follicles were excluded from further analysis if less than 20% of cells stained with CD20. To assess germinal center formation (secondary follicles), we scored the follicles using both structural and staining information; follicles received a score of 2 if more than 5% of cells in the follicle stained for BCL-6 and the follicle had a visible germinal center and mantle zone structure (secondary follicle), a score of 1 if either more than 5% of cells in the follicle stained for BCL6 or if the follicle had a visible germinal center and mantle zone structure (secondary follicle), and a score of 0 if there was not staining for BCL6 in above 5% of the cells in the follicle nor did the follicle have a visible germinal center and mantle zone structure (primary follicle).

## Spatial Transcriptomics

### Visium CytAssist

Visium CytAssist Spatial Gene Expression assay was performed following manufacturer’s (10x Genomics) instructions. Briefly, RNA quality was assessed using the Agilent TapeStation, and lymph node samples with a DV200 ≥30% were selected for analysis. FFPE tissue sections that were placed on a plain glass slide were deparaffinized and decrosslinked. Human whole transcriptome probe panels consisting of specific probes for each targeted gene were then added to the tissue. These probe pairs hybridize to their gene target and were then ligated to one another. Tissue slides and Visium CytAssist Spatial Gene Expression Slides were loaded into the Visium CytAssist instrument, where they were brought into proximity with one another. Each Visium CytAssist Slide contains about 5,000 Capture Areas (55 um across) with barcoded spots that include oligonucleotides required to capture gene expression probes. Gene expression probes were released from the tissue upon CytAssist Enabled RNA Digestion & Tissue Removal, enabling capture by the spatially barcoded oligonucleotides present on the Visium slide surface. All the probes captured on a specific spot share a common Spatial Barcode. Libraries were generated from the probes and sequenced, and the Spatial Barcodes were used to associate the reads back to the tissue section images for spatial mapping of gene expression. Sequencing was performed on an Illumina NovaSeq X sequencer with depth set at 25,000 read per capture spot. FASTQ files were subsequently loaded on to Space Ranger (10x Genomics) for read alignment, spot detection, gene expression quantification and data visualization.

### Gene expression analysis

The Space Ranger output (filtered_feature_bc_matrix.h5) was loaded into R (v 4.3.1) using Seurat (v 4.4.0), with a total of 25,750 capture spots across all 6 lymph nodes (13,556 D21 capture spots and 12,194 T21 capture spots). Filtering was performed, removing any spots with < 200 genes per spot (nFeature_Spatial < 200) and a mitochondrial percentage < 10% (the percentage of mitochondrial gene usage per spot), resulting in 24,678 high quality capture spots (13,502 D21 spots, 11,176 T21 capture spots). Normalization was run using Seurat SCTransform^47^, on each sample individually. Harmony integration was performed after normalization and PCA, with group (D21 vs T21) as the batch variable. Clustering and UMAP analysis were conducted via FindNeighbors, FindClusters (with a resolution of 0.3), and RunUMAP. To assess differential gene expression within each cluster, SCTransform values were prepped using PrepSCTFindMarkers and FindAllMarkers was run.

### Cluster assignment

We performed cluster annotation via examining the expression of a set of marker genes for different lymph node tissue regions, as well as by examining the regions identified via immunohistochemistry staining. The following marker genes were examined: *CXCL13, CD79A,* and *CD79B* for B cell follicles; *CCR7, CCL19, CCL21, IL7R, TRBC1,* and *TRCB2* for T cell zones/paracortex; *LYVE1, CD209, MARCO, CLEC4M, IGKC, IGHG1,* and *IGHA1* for medulla clusters; *COL1A1, COL1A2, COL3A1, NTS, TNFRSF9, CXCL5,* and *ACKR1* for the subcapsular sinus; *HBD*, *HBA2*, and *HBB* for red blood cell infiltration; and *MYH11, MCAM, ACTA2*, *COL1A1, COL1A2,* and *COL3A1* for the capsule and surrounding muscle tissue. The medulla clusters were separated out by higher expression of stromal markers (*LYVE1, CD209, MARCO, CLEC4M*; medullary sinus (medulla #1)), higher expression of B cell markers (*IGKC, IGHG1,* and *IGHA1;* medullary cords (medulla #2)), expression of neutrophil markers (*S100A8, S100A9, MPO, AZU1, CTSG*; medulla with neutrophil infiltration (medulla #3)), and a general medulla cluster which represented few spots across the lymph nodes (medulla #4).

### Calculation of pseudobulk estimates

We summarized spot expression values into bulk-like expression estimates for downstream gene expression modeling. For each tissue type cluster per sample, raw UMI counts were summed across all spots assigned to that sample for each gene using the sparse_Sums function in textTinyR (v1.1.8)^48^, yielding an n x m expression matrix for each cluster, where n is the number of samples included in the study and m is the number of genes detected in the spatial transcriptomics analysis.

### Calculation of residuals for gene expression modeling

For each tissue type (cluster), lowly-expressed genes were removed if logCPM < 2, retaining the following number of genes: cluster 0 (Medullary sinus (Medulla #1)) = 17437, cluster 5 (Medullary cords (Medulla #2)) = 17247, cluster 7 (Medulla with neutrophil infiltration (Medulla #3)) = 17511. After removing lowly-expressed genes, normalization factors to scale the raw library sizes were calculated using calcNormFactors in edgeR (v 4.0.16)^49^. The voom function in limma (v3.58.1)^50^ was used to apply these size factors, estimate the mean-variance relationship, and convert raw pseudocounts to logCPM values.

### Modeling the effect of T21 on gene expression

The following linear model was used to identify genes with differential expression levels between D21 and T21 lymph node clusters:

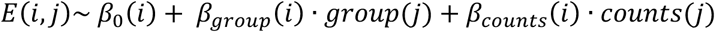

Here, *E*(*i*, *j*) represents the dataset-corrected expression estimate of gene i for individual j, *β*_0_(*i*)is the global intercept, and *β*_g*roup*_(*i*) represents the effect of the group (D21 vs T21) on gene i’s expression. Counts represents the number of genes (nCount_Spatial) captured within that cluster type for sample j, accounting for differences in spot number and gene number between samples, with *β_counts_*(*i*) capturing the effect of nCount_Spatial on expression. The model was fit using the lmFit and eBayes functions in limma, and the estimates of *β*_g*roup*_(*i*) were extracted with their corresponding p-values (adjusted using the Benjamini-Hochberg correction). Genes were graphed if they had an absolute *log*_2_ fold change of at least 1.5 and an adjusted p-value of less than 0.1.

## 10× Genomics paired single-cell RNA-sequencing and V(D)J sequencing

### Cell sorting

Donor PBMCs were thawed in complete RPMI and rested overnight. PBMCs were stained with Live/Dead Zombie Aqua (1:1000 in PBS), then FcBlock (1:20), APC-Cy7 CD19 (1:100), and PE-Cy7 CD4 (1:100) in FACS buffer (0.1% BSA + PBS). 5000 – 30,000 CD19+ B cells were sort-purified using a Bigfoot Spectral Cell Sorter (Invitrogen) and brought directly to the Columbia Genomics Core for library preparation and sequencing.

### Single-cell RNA-seq and V(D)J library preparation and sequencing

Sequencing libraries were prepared using the Chromium GEM-X Single Cell 5’ Reagent Kit (v3, 10x Genomics) and the Chromium Single Cell V(D)J BCR Amplification Kit (10x Genomics) according to manufacturer’s instructions. Libraries were sequenced on a NovaSeq X.

### Data processing

Gene expression FASTQ files from each sample library were mapped to a GRCh38 human reference and V(D)J FASTQ files were mapped to a V(D)J human reference (Human reference (GRCh38) - 2020-A and 5.0 V(D)J human reference, respectively, downloaded from 10x Genomics), using the “multi” pipeline in Cell Ranger (v 10.0.0)^51^. Cell-level quality control filtering was performed using Seurat (v.5.5.0, R v.4.5.3)^52^. Cells were retained for downstream analysis if they had between 500 and 4000 genes detected (nFeature_RNA), between 2000 and 25000 total molecules detected (nCount_RNA), and if mitochondrial UMI usage was less than 20%, resulting in 90,782 high quality cells.

### Gene expression analysis

Gene filtering was performed using CreateSeuratObject min.cells, in which only genes present in at least 5 cells were kept. Normalization was performed on each sample (using NormalizeData, FindVariableFeatures, and ScaleData)^52^, regressing out a variable corresponding to the percent of mitochondrial UMIs per cell. PCA was conducted on each sample (RunPCA function) and then samples were integrated using Harmony integration^28^, via IntegrateLayers. After integration, the nearest neighbors were calculated (FindNeighbors function, dims = 1:30, all other parameters default) and cell clusters were called (FindClusters function, resolution = 0.2, all other parameters default). Dimensionality reduction was performed using UMAP (RunUMAP function, dims = 1:30).^52^ A small population was filtered out with a UMAP_1 score over 10, and the subsequent clusters were then analyzed for cell type.

### Cell type assignment

We performed cell type annotation via examining the expression of a set of marker genes for different B cell subsets (Figure 4B). The following marker genes were examined: *BACH2, TCL1A, IL4R, IGHD,* and *IGHM* for naïve B cells; *STAT1, MX1, ISG15,* and *IFI44L* for ISG+ naïve B cells; *CD69, MYC,* and *FOS* for activated naïve B cells; *CXCR3, ITGAX, FCLR5,* and *ZEB2* for CXCR3+ atypical B cells; *MIR155HG, NME1,* and *TNFRSF18* for miR-155+ B cells; *CD27, AIM2, CD86, IGHA1, IGHG1,* and *IGHG2* for memory and CD86+ switched B cells; and *PRDM1, XBP1, JCHAIN,* and *MZB1* for antibody-secreting cells. Cells were also assigned a gene set score (for genes *ITGAX, FCLR5,* and *ZEB2*) using Seurat AddModuleScore and reassigned to the CXCR3+ atypical B cell cluster if they scored over a 0.75. To further confirm the B cell subset identities, we assessed their corresponding heavy chain BCRs for SHM frequency (Supplemental Figure 5A) and isotype usage (Supplemental Figure 4B), which were consistent with expected values.

### Calculation of pathway enrichment score

To calculate module scores, we used the R package escape (v2.6.2)^53^, which integrates functions from the R package UCell (v2.14.0)^40^, a gene signature scoring method optimized for single-cell expression data. To compute module scores, we used the pre-defined MSigDB hallmark gene sets^54^ and the escape.matrix function (method = “UCell”) on the processed and labeled GEX Seurat object.

### V(D)J analysis

After CellRanger processing, AIRR compliant rearrangement data was generated from 10x V(D)J FASTA files using IgBlast^55^ via Change-O (v. 1.3.4)^56^ scripts AssignGenes.py and MakeDb.py. The resulting tsv file for each sample was modified to append a column with the sample ID and to append the sample ID to the cell barcodes; all samples were then merged into a single file. Inconsistent and nonproductive sequenced were filtered out, and only cells with a single heavy chain sequence paired with at least one light chain sequence were retained, as per the Immcantation analysis pipeline. The processed and labeled gene expression Seurat object was loaded and GEX and BCR cell ids were matched, cluster annotations transferred to the BCR data, and cells without GEX data were removed. Clonal analysis was performed using SHazaM (v. 1.3.2)^56^ and SCOPer (v. 1.5.0)^57^; the nearest-neighbor distance distribution was calculated (distToNearest function, split between samples and using only heavy chains) and the clonal threshold was calculated (findThreshold function). Clonal groups were defined based on junction region sequence similarity within partitions that share the same V gene, J gene, and junction length (hierarchicalClones^58^ function, per sample). Germline sequences were then reconstructed via dowser (v. 2.4.1)^59^ using the IGMT germline references^60^ (createGermlines function, per sample). Somatic hypermutation frequency was calculated using the observedMutations function from SHazaM, calculating frequency of combined replacement and silent mutations in the heavy chains.

### VH gene analysis

VH gene usage was calculated using the Alakazam (v.1.4.3)^56^ (countGenes function). We filtered out any VH genes that do not appear in at least one D21 sample and one T21 sample.

## Notes

### Author Declarations

The Icahn School of Medicine at Mount Sinai IRB gave ethical approval for this work. The Columbia University Irving Medical Center IRB gave ethical approval for this work.

## References

1 Mai, C. T. et al. National population-based estimates for major birth defects, 2010-2014. Birth Defects Res 111, 1420–1435 (2019). 10.1002/bdr2.1589

2 Bull, M. J. Down Syndrome. N Engl J Med 382, 2344–2352 (2020). 10.1056/NEJMra1706537

3 Antonarakis, S. E. et al. Down syndrome. Nat Rev Dis Primers 6, 9 (2020). 10.1038/s41572-019-0143-7

4 Pierce, M. J., LaFranchi, S. H. & Pinter, J. D. Characterization of Thyroid Abnormalities in a Large Cohort of Children with Down Syndrome . Horm Res Paediatr 87, 170–178 (2017). 10.1159/000457952

5 Fitzpatrick, V. et al. Prevalence of Infectious Diseases Among 6078 Individuals With Down Syndrome in the United States. J Patient Cent Res Rev 9, 64–69 (2022). 10.17294/2330-0698.1876

6 Malle, L. et al. Excessive negative regulation of type I interferon disrupts viral control in individuals with Down syndrome. Immunity 55, 2074–2084.e2075 (2022). 10.1016/j.immuni.2022.09.007

7 Eijsvoogel, N. B. et al. Declining antibody levels after hepatitis B vaccination in Down syndrome: A need for booster vaccination? J Med Virol 89, 1682–1685 (2017). 10.1002/jmv.24813

8 Sali, M. et al. SARS-CoV-2 vaccine humoral response in adults with Down syndrome. Clin Microbiol Infect 28, 1155.e1151–1155.e1154 (2022). 10.1016/j.cmi.2022.04.008

9 Valentini, D. et al. Generation of switched memory B cells in response to vaccination in Down syndrome children and their siblings. Vaccine 33, 6689–6696 (2015). 10.1016/j.vaccine.2015.10.083

10 Kusters, M. A. et al. Impaired avidity maturation after tetanus toxoid booster in children with Down syndrome. Pediatr Infect Dis J 30, 357–359 (2011). 10.1097/INF.0b013e3181ff85a8

11 de Hingh, Y. C. et al. Intrinsic abnormalities of lymphocyte counts in children with down syndrome. J Pediatr 147, 744–747 (2005). 10.1016/j.jpeds.2005.07.022

12 Malle, L. et al. Autoimmunity in Down’s syndrome via cytokines, CD4 T cells and CD11c(+) B cells. Nature 615, 305–314 (2023). 10.1038/s41586-023-05736-y

13 Waugh, K. A. et al. Mass Cytometry Reveals Global Immune Remodeling with Multi-lineage Hypersensitivity to Type I Interferon in Down Syndrome. Cell Rep 29, 1893–1908.e1894 (2019). 10.1016/j.celrep.2019.10.038

14 Lambert, K., et al. Deep immune phenotyping reveals similarities between aging, Down syndrome, and autoimmunity. Sci Transl Med 14, eabi4888 (2022). 10.1126/scitranslmed.abi4888

15 Dieudonné, Y. et al. Immune Defect in Adults With Down Syndrome: Insights Into a Complex Issue. Front Immunol 11, 840 (2020). 10.3389/fimmu.2020.00840

16 Briney, B., Inderbitzin, A., Joyce, C. & Burton, D. R. Commonality despite exceptional diversity in the baseline human antibody repertoire. Nature 566, 393–397 (2019). 10.1038/s41586-019-0879-y

17 Carsetti, R. et al. Reduced numbers of switched memory B cells with high terminal differentiation potential in Down syndrome. Eur J Immunol 45, 903–914 (2015). 10.1002/eji.201445049

18 Klein, U. & Dalla-Favera, R. Germinal centres: role in B-cell physiology and malignancy. Nat Rev Immunol 8, 22–33 (2008). 10.1038/nri2217

19 Farroni, C. et al. Dysregulated miR-155 and miR-125b Are Related to Impaired B-cell Responses in Down Syndrome. Front Immunol 9, 2683 (2018). 10.3389/fimmu.2018.02683

20 Roozendaal, R., Mebius, R. E. & Kraal, G. The conduit system of the lymph node. Int Immunol 20, 1483–1487 (2008). 10.1093/intimm/dxn110

21 Lütge, M. et al. Fibroblastic reticular cells form reactive myeloid cell niches in human lymph nodes. Sci Immunol 10, eads6820 (2025). 10.1126/sciimmunol.ads6820

22 Arroz-Madeira, S., Bekkhus, T., Ulvmar, M. H. & Petrova, T. V. Lessons of Vascular Specialization From Secondary Lymphoid Organ Lymphatic Endothelial Cells. Circ Res 132, 1203–1225 (2023). 10.1161/circresaha.123.322136

23 Heesters, B. A., Myers, R. C. & Carroll, M. C. Follicular dendritic cells: dynamic antigen libraries. Nat Rev Immunol 14, 495–504 (2014). 10.1038/nri3689

24 Krishnamurty, A. T. & Turley, S. J. Lymph node stromal cells: cartographers of the immune system. Nat Immunol 21, 369–380 (2020). 10.1038/s41590-020-0635-3

25 Takeda, A. et al. Single-Cell Survey of Human Lymphatics Unveils Marked Endothelial Cell Heterogeneity and Mechanisms of Homing for Neutrophils. Immunity 51, 561–572.e565 (2019). 10.1016/j.immuni.2019.06.027

26 Xiang, M. et al. A Single-Cell Transcriptional Roadmap of the Mouse and Human Lymph Node Lymphatic Vasculature. Front Cardiovasc Med 7, 52 (2020). 10.3389/fcvm.2020.00052

27 Heesters, B. A., van der Poel, C. E., Das, A. & Carroll, M. C. Antigen Presentation to B Cells. Trends Immunol 37, 844–854 (2016). 10.1016/j.it.2016.10.003

28 Korsunsky, I. et al. Fast, sensitive and accurate integration of single-cell data with Harmony. Nat Methods 16, 1289–1296 (2019). 10.1038/s41592-019-0619-0

29 Hao, Y. et al. Integrated analysis of multimodal single-cell data. Cell 184, 3573–3587.e3529 (2021). 10.1016/j.cell.2021.04.048

30 Dugan, H. L. et al. Profiling B cell immunodominance after SARS-CoV-2 infection reveals antibody evolution to non-neutralizing viral targets. Immunity 54, 1290–1303.e1297 (2021). 10.1016/j.immuni.2021.05.001

31 Wang, Y. et al. Integrating single-cell RNA and T cell/B cell receptor sequencing with mass cytometry reveals dynamic trajectories of human peripheral immune cells from birth to old age. Nat Immunol 26, 308–322 (2025). 10.1038/s41590-024-02059-6

32 Gong, Q. et al. Multi-omic profiling reveals age-related immune dynamics in healthy adults. Nature 648, 696–706 (2025). 10.1038/s41586-025-09686-5

33 Gao, X. et al. Zeb2 drives the formation of CD11c(+) atypical B cells to sustain germinal centers that control persistent infection. Sci Immunol 9, eadj4748 (2024). 10.1126/sciimmunol.adj4748

34 Tangye, S. G. Human CD21(lo)T-bet(+) B cells: Not as easy as “ABC”! J Hum Immun 2, e20260023 (2026). 10.70962/jhi.20260023

35 Espinoza, D. A. et al. Distinct stage-specific transcriptional states of B cells derived from human tonsillar tissue. JCI Insight 8 (2023). 10.1172/jci.insight.155199

36 King, H. W. et al. Single-cell analysis of human B cell maturation predicts how antibody class switching shapes selection dynamics. Sci Immunol 6 (2021). 10.1126/sciimmunol.abe6291

37 Vigorito, E. et al. microRNA-155 regulates the generation of immunoglobulin class-switched plasma cells. Immunity 27, 847–859 (2007). 10.1016/j.immuni.2007.10.009

38 Zheng, S. et al. NME proteins regulate class switch recombination. FEBS Lett 593, 80–87 (2019). 10.1002/1873-3468.13290

39 Ramba, M. & Bogunovic, D. The immune system in Down Syndrome: Autoimmunity and severe infections. Immunol Rev (2023). 10.1111/imr.13296

40 Andreatta, M. & Carmona, S. J. UCell: Robust and scalable single-cell gene signature scoring. Comput Struct Biotechnol J 19, 3796–3798 (2021). 10.1016/j.csbj.2021.06.043

41 Zhang, H., Wang, L. & Chu, Y. Reactive oxygen species: The signal regulator of B cell. Free Radic Biol Med 142, 16–22 (2019). 10.1016/j.freeradbiomed.2019.06.004

42 Limon, J. J. & Fruman, D. A. Akt and mTOR in B Cell Activation and Differentiation. Front Immunol 3, 228 (2012). 10.3389/fimmu.2012.00228

43 Singh, A., Joshi, V., Jindal, A. K., Mathew, B. & Rawat, A. An updated review on activated PI3 kinase delta syndrome (APDS). Genes Dis 7, 67–74 (2020). 10.1016/j.gendis.2019.09.015

44 Bashford-Rogers, R. J. M. et al. Analysis of the B cell receptor repertoire in six immune-mediated diseases. Nature 574, 122–126 (2019). 10.1038/s41586-019-1595-3

45 Landes, S. D., Stevens, J. D. & Turk, M. A. Cause of death in adults with Down syndrome in the United States. Disabil Health J 13, 100947 (2020). 10.1016/j.dhjo.2020.100947

46 Matsumoto, R. et al. Induction of bronchus-associated lymphoid tissue is an early life adaptation for promoting human B cell immunity. Nat Immunol 24, 1370–1381 (2023). 10.1038/s41590-023-01557-3

47 Hafemeister, C. & Satija, R. Normalization and variance stabilization of single-cell RNA-seq data using regularized negative binomial regression. Genome Biol 20, 296 (2019). 10.1186/s13059-019-1874-1

48 textTinyR: Text Processing for Small or Big Data Files (2023).

49 Chen, Y., Chen, L., Lun, A. T. L., Baldoni, P. L. & Smyth, G. K. edgeR v4: powerful differential analysis of sequencing data with expanded functionality and improved support for small counts and larger datasets. Nucleic Acids Res 53 (2025). 10.1093/nar/gkaf018

50 Ritchie, M. E. et al. limma powers differential expression analyses for RNA-sequencing and microarray studies. Nucleic Acids Res 43, e47 (2015). 10.1093/nar/gkv007

51 Zheng, G. X. et al. Massively parallel digital transcriptional profiling of single cells. Nat Commun 8, 14049 (2017). 10.1038/ncomms14049

52 Stuart, T. et al. Comprehensive Integration of Single-Cell Data. Cell 177, 1888–1902.e1821 (2019). 10.1016/j.cell.2019.05.031

53 Borcherding, N. et al. Mapping the immune environment in clear cell renal carcinoma by single-cell genomics. Commun Biol 4, 122 (2021). 10.1038/s42003-020-01625-6

54 Liberzon, A. et al. The Molecular Signatures Database (MSigDB) hallmark gene set collection. Cell Syst 1, 417–425 (2015). 10.1016/j.cels.2015.12.004

55 Ye, J., Ma, N., Madden, T. L. & Ostell, J. M. IgBLAST: an immunoglobulin variable domain sequence analysis tool. Nucleic Acids Res 41, W34–40 (2013). 10.1093/nar/gkt382

56 Gupta, N. T. et al. Change-O: a toolkit for analyzing large-scale B cell immunoglobulin repertoire sequencing data. Bioinformatics 31, 3356–3358 (2015). 10.1093/bioinformatics/btv359

57 Nouri, N. & Kleinstein, S. H. A spectral clustering-based method for identifying clones from high-throughput B cell repertoire sequencing data. Bioinformatics 34, i341–i349 (2018). 10.1093/bioinformatics/bty235

58 Gupta, N. T. et al. Hierarchical Clustering Can Identify B Cell Clones with High Confidence in Ig Repertoire Sequencing Data. J Immunol 198, 2489–2499 (2017). 10.4049/jimmunol.1601850

59 Hoehn, K. B., Pybus, O. G. & Kleinstein, S. H. Phylogenetic analysis of migration, differentiation, and class switching in B cells. PLoS Comput Biol 18, e1009885 (2022). 10.1371/journal.pcbi.1009885

60 Lefranc, M. P. et al. IMGT®, the international ImMunoGeneTics information system® 25 years on. Nucleic Acids Res 43, D413–422 (2015). 10.1093/nar/gku1056

